# Comparison of OneChoice® AI-based clinical decision support recommendations with infectious disease specialists and non-specialists for bacteremia treatment in Lima, Peru

**DOI:** 10.1101/2025.08.15.25333753

**Authors:** Juan Carlos Gómez de la Torre, Ari Frenkel, Carlos Chavez-Lencinas, Alicia Rendon, Max Fabian, José Caceres, Miguel Hueda-Zavaleta

## Abstract

Bacteremia is a major contributor to global morbidity and mortality, particularly in low- and middle-income countries where diagnostic delays and empirical antimicrobial misuse exacerbate resistance. This study assessed the accuracy of OneChoice®, an artificial intelligence (AI)-based Clinical Decision Support System (CDSS), in guiding antimicrobial therapy for bloodstream infections (BSIs) in Lima, Peru. A retrospective, observational design was used, comparing therapeutic recommendations generated by OneChoice®—based on molecular (FilmArray®) and phenotypic (MALDI-TOF MS, VITEK2) data—with the clinical decisions of 94 physicians (35 infectious disease [ID] specialists and 59 non-specialists) across 366 survey-based evaluations of bacteremia cases. Concordance between CDSS and physician decisions was analyzed using Cohen’s Kappa and logistic regression. The overall concordance rate was 96.14% when considering any suggested treatment, and 74.59% for the top recommendation, with a substantial agreement (κ = 0.70). ID specialists showed significantly higher concordance (κ = 0.78) than non-ID physicians (κ = 0.61), and specialization was the strongest predictor of agreement (OR = 2.26, p = 0.001). Escherichia coli cases had the highest concordance, while Pseudomonas aeruginosa showed the lowest. The CDSS reduced inappropriate antibiotic use, particularly unnecessary carbapenem prescriptions. These findings support the utility of AI-CDSS tools in enhancing antimicrobial stewardship and standardizing care, especially in resource-limited healthcare settings.

## 1. Introduction

Sepsis and bloodstream infections (BSIs) represent a critical global health issue, particularly in low- and middle-income countries (LMICs), where delayed diagnoses and treatment gaps compound mortality [1]. In 2020 alone, an estimated 11 million deaths globally were attributed to sepsis, making it one of the leading causes of death worldwide [2]. In Latin America, limited diagnostic capacity and inappropriate antibiotic use exacerbate the crisis, leading to increasing multidrug-resistant (MDR) infections [2].

Rapid and accurate diagnosis is essential for effective management, as delays in appropriate antimicrobial therapy have been associated with increased mortality [3]. Traditional diagnostic methods, such as blood cultures and phenotypic antimicrobial susceptibility testing (AST), remain the gold standard but suffer from long turnaround times, typically requiring 24 to 72 h for actionable results. This diagnostic delay is directly associated with increased mortality and inappropriate empirical therapy [4].

BSIs caused by organisms like *Escherichia coli*, *Klebsiella pneumoniae*, and *Pseudomonas aeruginosa* are increasingly resistant to standard antibiotics, driving the need for faster and more precise diagnostic tools [5]. Recent advances in molecular diagnostics, including FilmArray^®^ (BioFire Diagnostics, LLC, Salt Lake City, UT, USA) and GeneXpert^®^ (Cepheid, Sunnyvale, CA, USA), have significantly reduced this time from days to hours, allowing for faster pathogen identification and resistance profiling [6]. However, effectively integrating such diagnostic tools into clinical workflows remains challenging, necessitating more robust decision support mechanisms.

When integrated with artificial intelligence (AI), these tools form the backbone of clinical decision support systems (CDSSs) and can provide personalized antimicrobial recommendations [7]. AI-based CDSSs, such as OneChoice^®^ developed by Arkstone, are particularly promising for infectious disease management because they synthesize vast datasets in real time and align with clinical guidelines. This AI-CDSS integrates molecular (e.g., FilmArray) and phenotypic (e.g., VITEK2 and MALDI-TOF MS) diagnostic outputs to offer antimicrobial treatment recommendations. Its design aligns with global and regional stewardship goals, including those from the Surviving Sepsis Campaign and IDSA [8,9]. The OneChoice^®^ CDSS, powered by a machine learning model, has demonstrated strong internal validity in antimicrobial stewardship. In one study that examined the accuracy of its internal validation, the model revealed that it can accurately distinguish trained from novel data (100% accuracy) and showed 84% agreement with clinical standards in minor discrepancies while avoiding major discrepancies entirely. These results, confirmed through multiple validation methods and enhanced by human-in-the-loop (HITL) oversight, support the system’s potential as a reliable tool for optimizing antimicrobial therapy [10].

These systems have demonstrated potential in reducing inappropriate antibiotic use, optimizing dosing, and ensuring compliance with antimicrobial stewardship principles [11]. While initial results show promise in high concordance with expert ID physicians’ recommendations, real-world validation remains limited, especially in settings such as Peru.

Artificial intelligence (AI) and machine learning (ML) advancements have revolutionized diagnostics in infectious diseases. AI models are increasingly integrated into CDSSs, providing real-time, tailored therapeutic recommendations based on molecular and phenotypic data [11]. Such systems have effectively reduced diagnostic turnaround times and improved clinical outcomes [12]. AI-CDSS applications include early sepsis detection, resistance prediction, and antimicrobial therapy optimization [13,14]. Moreover, in regions like Latin America, AI-driven CDSSs may help bridge technological gaps and support overburdened healthcare systems [15].

Given the complex interplay of clinical, microbiological, and pharmacological factors in BSI management, AI-driven CDSSs have the potential to standardize therapeutic approaches and minimize errors due to human variability. AI-enhanced platforms can analyze diverse datasets in real time, offering context-specific recommendations that align with stewardship principles [16]. OneChoice^®^ is designed to mitigate the therapeutic lag between pathogen detection and treatment initiation, particularly critical in time-sensitive infections such as sepsis. OneChoice^®^ has demonstrated that it can deliver rapid and accurate therapeutic recommendations in cases of bacteremia. An 80% concordance was observed between the recommendations generated from molecular and phenotypic results. Furthermore, molecular-based recommendations were available up to 29 h earlier than those based on phenotypic data, suggesting that integrating molecular diagnostics with AI-driven CDSSs could significantly accelerate clinical decision making in bacteremia. Notably, most discrepancies were due to the inability of certain molecular platforms to detect specific phenotypic resistance mechanisms [17].

Despite the promise of CDSS platforms, several barriers persist. These include clinician skepticism, system interoperability issues, and a lack of real-world validation [12,18]. Additionally, implementation in LMICs like Peru is hindered by limited infrastructure and low CDSS awareness among healthcare professionals [15].

Studies evaluating CDSSs such as OneChoice^®^ remain limited, and few investigations have rigorously assessed its alignment with expert clinical judgment in LMICs, particularly among non-specialist physicians.

In Peru, a country with documented high resistance rates and an underfunded infectious disease infrastructure [15], the clinical validation of AI-CDSS tools is critically needed. This study is among the first to evaluate the agreement between an AI-CDSS and physicians across specialties, providing crucial insights into how AI can bridge knowledge gaps and enhance precision medicine. This study compares the CDSS recommendations with therapeutic decisions made by infectious disease (ID) specialists and non-ID specialists, assessing their concordance. The findings will help inform implementation frameworks, training needs, and policy recommendations for AI’s ethical and scalable deployment in infectious disease care.

## 2. Materials and Methods

### Study Design and Setting

This study employed a retrospective, observational design, conducted in tertiary healthcare facilities in Lima, Peru. The study utilized molecular and phenotypic diagnostic methods routinely applied to guide antimicrobial therapy. The primary objective was to compare the CDSS recommendations with therapeutic decisions made by infectious disease (ID) specialists and non-ID specialists, assessing their concordance.

### Participants and Selection Criteria

The study involved 94 physicians, comprising 35 infectious disease specialists and 59 non-ID specialists. Participants were selected based on their involvement in bacteremia management within the healthcare facilities. The inclusion criteria mandated that participating physicians had at least three years of clinical experience in infectious disease management. The study included confirmed bacteremia cases, selected based on strict inclusion criteria to ensure the reliability of the dataset. Eligible cases involved patients with positive blood cultures confirmed through FilmArray® molecular testing and phenotypic identification using MALDI-TOF MS and VITEK2 systems. Cases were excluded if they had incomplete laboratory data, invalid susceptibility results, or were determined to be contaminants rather than true BSIs.

### Data Acquisition and Characteristics

Data for the study were acquired from confirmed bacteremia cases, integrating microbiological, clinical, and therapeutic information. The dataset included results from molecular diagnostics using the FilmArray Blood Culture Identification (BCID) Panel (BioFire Diagnostics, LLC, Salt Lake City, UT, USA) and phenotypic antimicrobial susceptibility tests conducted with Matrix-assisted laser desorption ionisation time-of-flight (MALDI-TOF MS) (bioMérieux, Inc. France) and VITEK2 system (bioMérieux, Inc. France). The initial OneChoice® report (**<u>Supplement 1</u>**) was generated based on data from molecular results alone, while the final OneChoice® (Fusion) report (**<u>Supplement 2</u>**) incorporated both molecular and phenotypic test results.

The study focused on bacterial BSIs, selecting cases with available molecular and phenotypic testing results. Clinical case surveys were administered to physicians, presenting cases twice, initially with molecular results alone, and later with combined molecular and phenotypic data. Responses were collected to evaluate the agreement between physicians and OneChoice®. In total, 366 case evaluations were collected, allowing for a direct comparison of CDSS recommendations versus clinical judgment (**<u>Supplement 3</u>**).

### Study Procedures and Tools/Instruments/Materials/Equipment

The experimental procedures followed a structured approach to ensure consistency and reproducibility. Blood samples from patients with suspected bacteremia were processed using the FilmArray BCID Panel, which operates under specific conditions: samples were incubated at 37°C for 24 hours before analysis. The MALDI-TOF MS was calibrated daily using standard bacterial strains to ensure accuracy in phenotypic identification. The VITEK2 system was employed for susceptibility testing, using standard inoculum sizes and incubation periods as per manufacturer guidelines.

Each physician received clinical case surveys electronically, with cases presented randomly to minimize bias. Surveys were designed using Microsoft Forms, ensuring uniformity in data collection (**<u>Supplement 3</u>**). The CDSS utilized proprietary algorithms to generate therapeutic recommendations, which were then compared to those of physicians.

### Data Preparation

Before analysis, all collected data were anonymized to protect patient confidentiality. Data entry was performed using Microsoft Excel, with each case assigned a unique numeric code. The dataset underwent rigorous cleaning procedures to ensure accuracy, including validation checks for duplicate entries and cross-referencing against laboratory records to confirm diagnostic results (**<u>Supplement 4</u>**).

In this study, concordance was defined as agreement between the therapeutic recommendation provided by the CDSS and the expert review based on molecular and/or phenotypic data by the Infectious and Tropical Disease and Non-Infectious Disease Specialists. Discordance refers to any disagreement with expert judgment regarding antimicrobial selection, dosing, or spectrum. Discordant cases were further categorized by type of mismatch, including incorrect antibiotic choice; incorrect dosing or interval; both incorrect antibiotic choice and dosing; unnecessary broad-spectrum coverage; unnecessary broad-spectrum with carbapenem overuse; or a combination of these. Multidrug-resistant (MDR) organisms were defined as non-susceptible to at least one agent in three or more antimicrobial categories, while extensively drug-resistant (XDR) organisms were non-susceptible to all but one or two categories, according to international consensus definitions [19].

### Data Analysis

Statistical analyses were conducted using STATA v.17. Descriptive statistics summarized physician responses, concordance rates, and frequency distributions of bacterial species and resistance genes. Cohen’s Kappa coefficient (κ) assessed inter-rater agreement, with interpretations based on established scales for agreement levels. Chi-square (χ²) tests examined associations between categorical variables, such as specialty, experience level, and concordance rates. Fisher’s exact test was applied for bacteria-associated bacteremia cases with limited sample sizes.

Multivariate logistic regression identified independent predictors of concordance, adjusting for specialty, experience, and antimicrobial resistance status. Model specifications included interaction terms to explore the influence of combined factors on therapeutic decision-making.

To assess the concordance between the CDSS and the clinical judgment of the participants, each therapeutic recommendation generated by the CDSS was independently reviewed by two infectious disease specialists (JCGdlT and CChL). Cases were classified as concordant or discordant depending on whether the participant’s decision aligned with the CDSS recommendation. For discordant cases, an adjudication process was conducted to determine whether the discrepancy favored the CDSS, the participant, or both. When the two infectious disease specialists disagreed in their classifications, a third specialist (MH-Z) evaluated the case, and a consensus decision was reached. In addition, all discrepancies were categorized into five mutually exclusive groups: incorrect antibiotic choice; combined errors in antibiotic selection and dosing or duration; unnecessary use of broad-spectrum carbapenems; unnecessary use of broad-spectrum antibiotics only; and discrepancies attributable to both perspectives. The study was approved for ethical approval to the Institutional Ethics Committee of the Private University of Tacna, Peru (FACSA-CEI/093-06-2025). Given its retrospective nature, the need for informed consent was waived. All data were fully anonymized to protect patient privacy, with names replaced by unique numeric codes to prevent patient identification. The study adhered to international ethical standards, ensuring confidentiality, data integrity, and research reproducibility. The findings will be submitted for peer-reviewed publication and presented at scientific conferences to promote awareness of CDSS integration in antimicrobial stewardship.

## 3. Results

A total of 366 bacteremia cases were evaluated, of which 206 (56.3%) were reviewed by infectious disease (ID) specialists and 160 (43.7%) by non-infectious disease (non-ID) physicians. A significant difference was observed in years of experience between the two groups (p = 0.001); 71.2% of ID specialists had between 1 and 5 years of experience, whereas 52% of non-ID physicians had between 16 and 20 years. The most frequently isolated bacterial species were *Escherichia coli* (35.5%), *Klebsiella pneumoniae/Klebsiella aerogenes* (15.8%), and *Salmonella typhi/enterica* (13.7%). *Klebsiella spp*. and *Salmonella spp*. were more commonly observed in cases evaluated by ID specialists (65.6% and 64%, respectively). Additionally, *Serratia marcescens* was exclusively identified in cases reviewed by ID specialists. Regarding resistance genes, 59% of isolates showed no detectable resistance genes, with a significant difference between groups (p = 0.047). ID specialists more frequently evaluated bacteremia cases involving strains with *CTM-X* (61.3% vs. 38.7%) and *NDM* genes (60% vs. 40%). All cases with *VIM* beta-lactamase genes were reported in the non-ID group. Overall, 39.9% of isolates were classified as multidrug-resistant, with no significant differences between groups (p = 0.640) (Table 1).

**Table 1:** Characteristics of evaluated bacteremia cases by specialist background: Microbial profiles, resistance genes, and concordance rates.

| Variable | Bacteremia cases evaluated |  |  | p-value |
| --- | --- | --- | --- | --- |
|  | Total<br>(n= 366) | by |  |  |
|  |  | Non-ID (n= 160) | ID (n= 206) |  |
| Specialty Time (%) |  |  |  | 0.001<sup>a</sup> |
| - 1 - 5 years | 104 (28.42) | 30 (28.8) | 74 (71.2) |  |
| - 16 - 20 years | 50 (13.66) | 26 (52.0) | 24 (48.0) |  |
| - > 20 years | 212 (57.92) | 104 (49.1) | 108 (50.9) |  |
| Bacteria Type (%) |  |  |  | 0.063<sup>a</sup> |
| - *Escherichia coli* | 130 (35.52) | 66 (50.8) | 64 (49.2) |  |
| - *Klebsiella pneumoniae/ aerogenes* | 58 (15.84) | 20(34.4) | 38(65.6) |  |
| - *Salmonella spp.* | 50 (13.66) | 18(36) | 28(64.0) |  |
| - *Streptococcus sp.* | 34 (9.3) | 18(53) | 16(47.0) |  |
| - *Pseudomonas aeruginosa* | 26 (7.10) | 12 (46.2) | 14 (53.8) |  |
| - *Enterococcus faecalis* | 18 (4.92) | 6 (33.3) | 12 (66.7) |  |
| - *Staphylococcus haemolyticus* | 16 (4.37) | 6 (37.5) | 10 (62.5) |  |
| - *Enterobacter sp.* | 14 (3.82) | 8 (57.1) | 6 (42.9) |  |
| - *Klebsiella oxytoca* | 10 (2.73) | 6 (60.0) | 4 (40.0) |  |
| - *Serratia marcescens* | 10 (2.73) | 0 (0.0) | 10 (100.0) |  |
| Resistance genes (%) |  |  |  | 0.047<sup>a</sup> |
| - No genes detected | 216 (59.02) | 90 (41.7) | 126 (58.3) |  |
| - CTM-X | 62 (2.7) | 24 (38.7) | 38 (61.3) |  |
| - ESBL | 56 (16.94) | 30 (53.6) | 26 (46.4) |  |
| - MecA/C | 16 (4.37) | 6 (37.5) | 10 (62.5) |  |
| - VIM beta-lactamase | 6 (1.64) | 6 (100.0) | 0 (0.0) |  |
| - NDM | 10 (2.73) | 4 (40.0) | 6 (60.0) |  |
| Multidrug resistant (%) |  |  |  | 0.640<sup>a</sup> |
| - Multidrug resistant | 146 (39.89) | 66 (45.2) | 80 (54.8) |  |
| - Not Multidrug resistant | 220 (60.11) | 94 (42.7) | 126 (57.3) |  |
| Concordance (%) |  |  |  | 0.001<sup>a</sup> |
| - Disagreement | 93 (25.41) | 54 (58.1) | 39 (41.9) |  |
| - Agreement | 273 (74.59) | 106 (38.8) | 167 (61.2) |  |
| Mismatch Type (%) |  |  |  | 0.001<sup>a</sup> |
| - Incorrect Antibiotic Choice | 45 (48.39) | 29 (64.4) | 16 (35.6) |  |
| - Incorrect Antibiotic Choice, | 18 (19.35) | 7 (38.9) | 11 (61.1) |  |
| Incorrect Dosing or Interval |  |  |  |  |
| - Incorrect Dosing or Interval | 6 (6.45) | 5 (83.3) | 1 (16.7) |  |
| - Unnecessary Broad Spectrum | 8 (8.6) | 4 (50.0) | 4 (50.0) |  |
| - Carbapenem overuse | 10 (10.75) | 7 (70.0) | 3 (30.0) |  |
| - More than one mismatch | 6 (6.45) | 2 (33.3) | 4 (66.7) |  |

a: Chi-square; CTX-M: Cefotaximase-Munich; ESBL: extended spectrum beta-
lactamase; NDM: New Delhi metallo- $\beta$ -lactamases; VIM: Verona integron-
encoded metallo- $\beta$ -lactamases.

In terms of concordance between the system’s recommendations and expert evaluation, 74.6% of cases showed overall agreement, with a significantly higher rate in the ID group (61.2% vs. 38.8%, p = 0.001). Disagreement was more common among non-ID physicians (58.1% vs. 41.9%) (Table 1). Finally, A detailed assessment of discrepancies in therapeutic decisions revealed that 92.47% of discordant cases favored the CDSS recommendations, while 7.53% reflected consideration of both CDSS and physician input. The most frequent type of mismatch was incorrect antibiotic selection (48.4%), followed by combined errors in antibiotic choice, dosing, or interval (19.4%), and unnecessary broad-spectrum use (8.6%). Additionally, 10.75% of mismatches involved unnecessary carbapenem overuse, underscoring the importance of carbapenem stewardship in the context of resistance. Notably, most dosing-related errors occurred in the non-ID group (Table 1, Figure 1).

**Figure 1:**
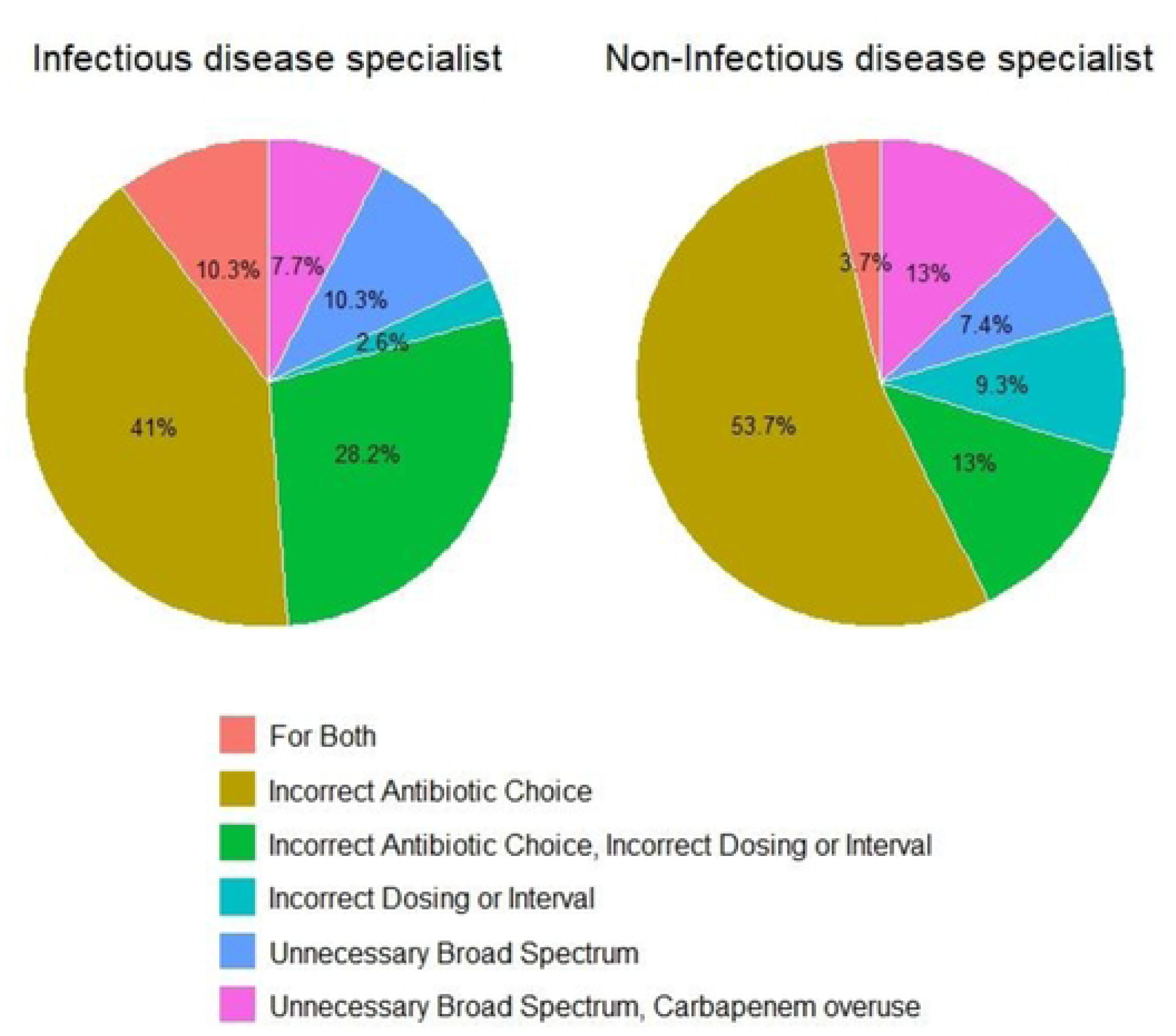
Discrepancy type between Infectious disease specialist and Non-Infectious Disease Specialist and result type

The overall concordance rate between OneChoice® recommendations and physician decisions was 96.14% when considering any of the four alternative treatment options provided by the system. Concordance was slightly lower when evaluating the best-recommended option, with rates of 74.32% for molecular-based results and 74.86% for phenotypic results. The Kappa Index, a statistical measure of agreement beyond chance, was calculated for all participants and stratified by specialty, the overall Kappa Index was 0.70, indicating substantial agreement. Infectious disease specialists demonstrated a higher concordance with a Kappa value of 0.78 than non-ID specialists, who had a Kappa value of 0.61 (Table 2).

**Table 2:**
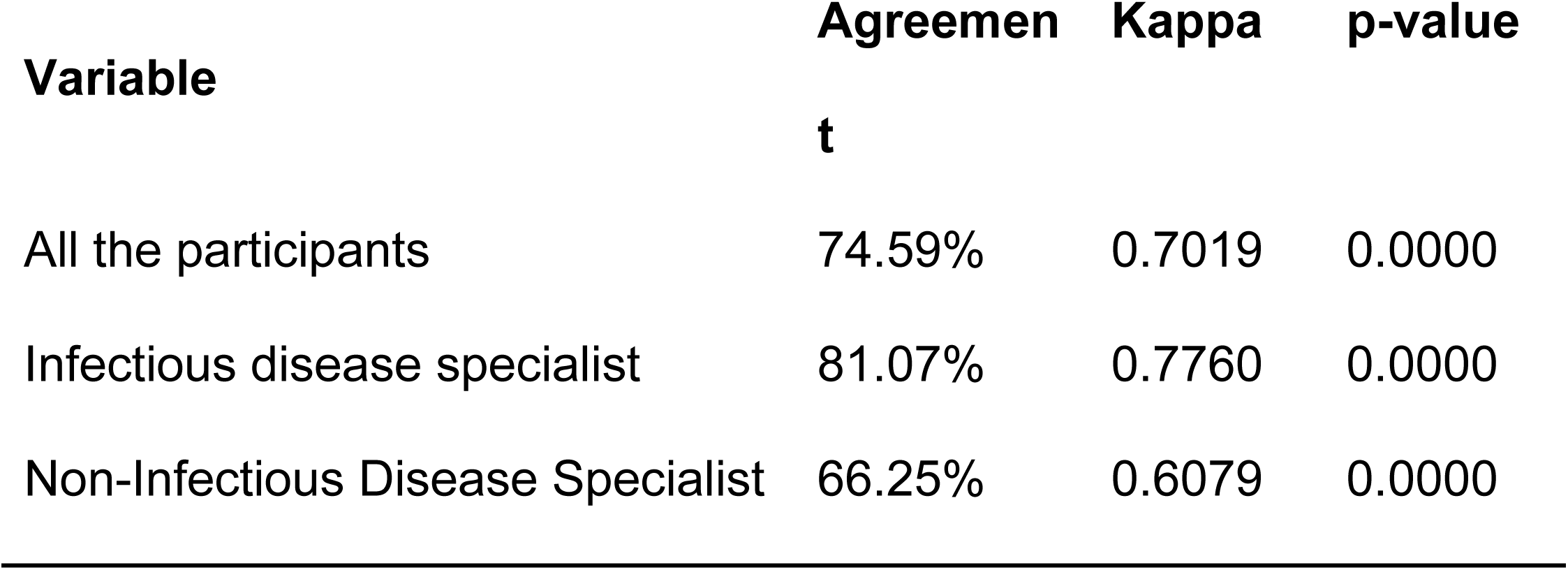
Kappa Index regarding Antibiotic prescription between SSDD and Physicians.

The study further explored concordance rates based on physician experience, revealing higher agreement among more experienced physicians, particularly among ID specialists. Concordance rates among ID specialists ranged from 70.83% for those with 16-20 years of experience to 84.26% for those with more than 20 years. In contrast, non-ID specialists showed an overall lower concordance rate of 66.25%, with the highest agreement observed in the 16–20-year experience group at 80.77% (Table 3). The analysis of bacterial species and concordance rates revealed that Escherichia coli had the highest concordance rate at 80%. In contrast, Pseudomonas aeruginosa exhibited the lowest agreement at 53.84%, with a statistically significant p-value of 0.012. Conversely, Staphylococcus haemolyticus showed 100% concordance (Table 3).

**Table 3:**
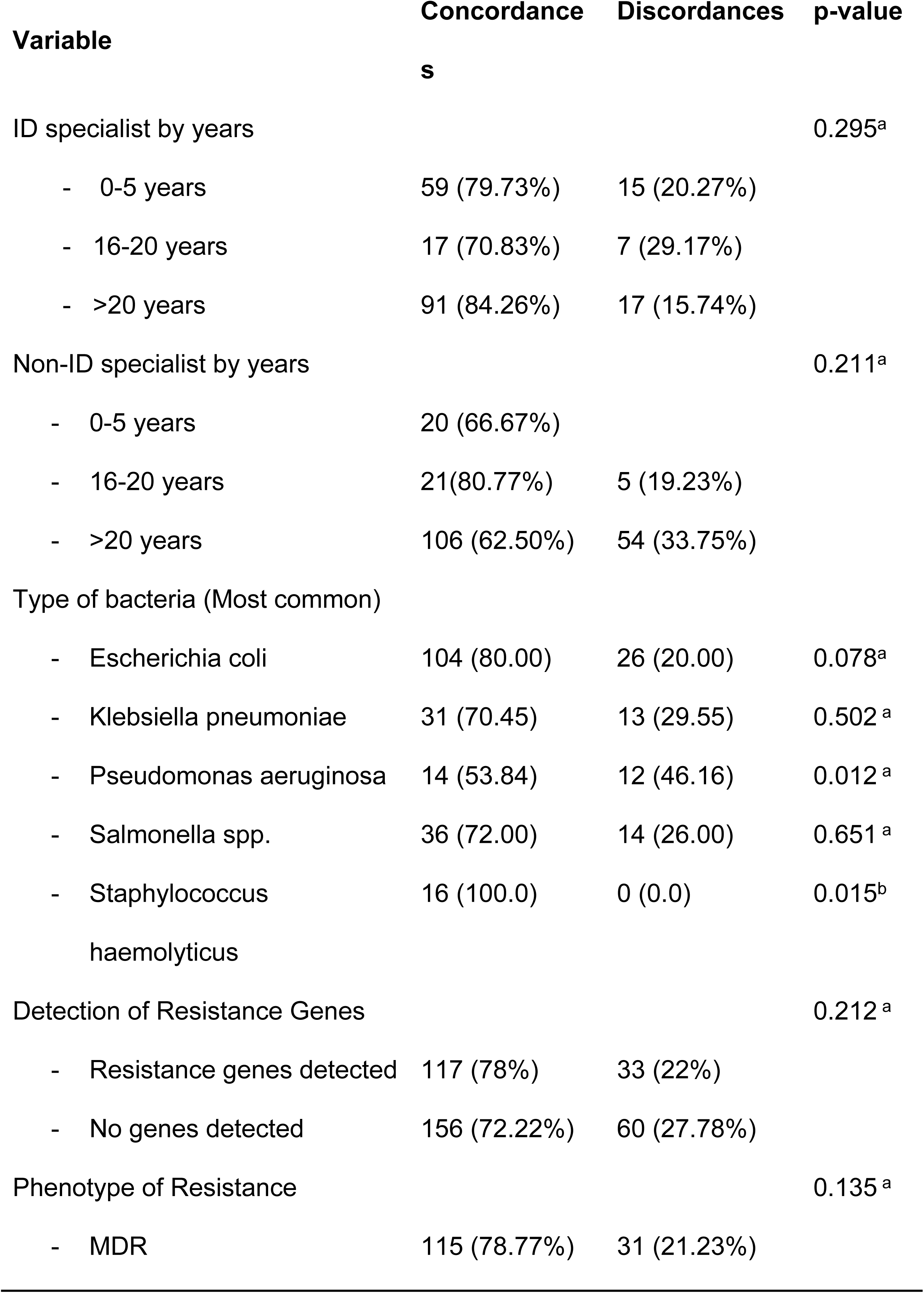
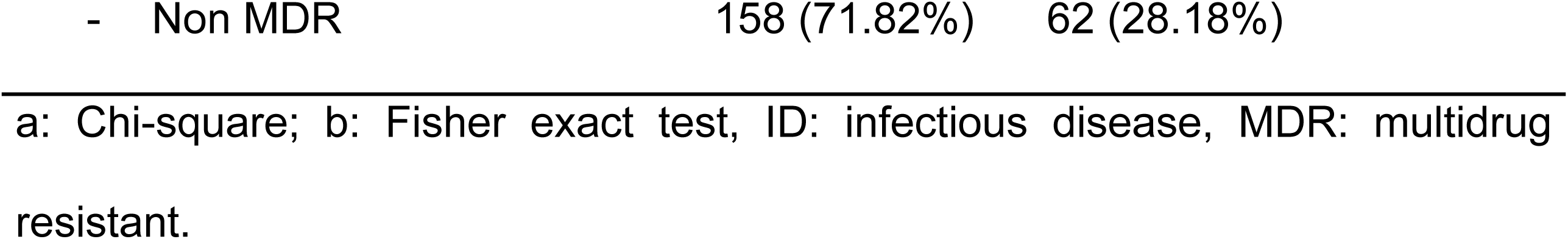
Frequencies of concordances and discordances in infectious disease and non-infectious disease doctors and type of discrepancies detected.

A multivariate logistic regression analysis was performed to identify factors influencing concordance, adjusting for specialty, years of experience, and MDR status. The results, summarized in Table 4, indicated that being an ID specialist was the strongest predictor of concordance, with an odds ratio (OR) of 2.26 and a p-value of 0.001. Years of experience did not show a statistically significant effect on concordance, with an OR of 1.27 (p = 0.562) for physicians with 16-20 years of experience and an OR of 1.06 (p = 0.843) for those with more than 20 years. MDR status showed a trend towards higher concordance (OR = 1.52, p = 0.104), suggesting that physicians are more likely to follow CDSS recommendations when dealing with multidrug-resistant infections. However, this result was not statistically significant (Table 4).

**Table 4:** Multivariate Logistic Regression Analysis of Factors Associated with Concordance Between Physician Decisions and OneChoice® Recommendations.

| Concordance | OR | p value | 95% CI |
| --- | --- | --- | --- |
| Specialty |  |  |  |
| - No infectious disease specialist |  | Reference |  |
| - Infectious disease specialist | 2.256 | **0.001** | 1.381 - 3.685 |
| Years of specialty |  |  |  |
| - 0 - 5 years |  | Reference |  |
| - 16-20 years | 1.272 | 0.562 | 0.563 - 2.871 |
| - > 20 years | 1.058 | 0.843 | 0.602 - 1.860 |
| Phenotype of Resistance |  |  |  |
| - Non MDR |  | Reference |  |
| - MDR | 1.517 | 0.104 | 0.9179 - 2.508 |

OR: odds ratio; CI: confidence interval; MDR: multidrug resistant.

Finally, we evaluated the overall level and nature of the discrepancies between the CDSS and the study participants. Concordance between the CDSS recommendations and the participants’ assessments occurred in 74.6% of cases (273/366), while the remaining 25.4% showed discordance (Figure 2a). Among the discordant cases, the vast majority (92.5%) favored the CDSS (Onechoice) recommendation, while only 7.5% were considered acceptable by both parties (CDSS and expert reviewers) (Figure 2b). Likewise, the most frequent types of discrepancies were incorrect antibiotic choice (51.7%), combined errors in antibiotic selection, dosing, or duration (20.7%), unnecessary use of broad-spectrum or carbapenem agents (11.5%), unnecessary use of broad-spectrum antibiotics (9.2%), and discrepancies attributable to both perspectives (6.9%) (Figure 2c). These findings suggest that most discrepancies were due to inappropriate antibiotic selection and that when disagreement occurred, the CDSS recommendations were more accurate in most cases.

**Figure 2.**
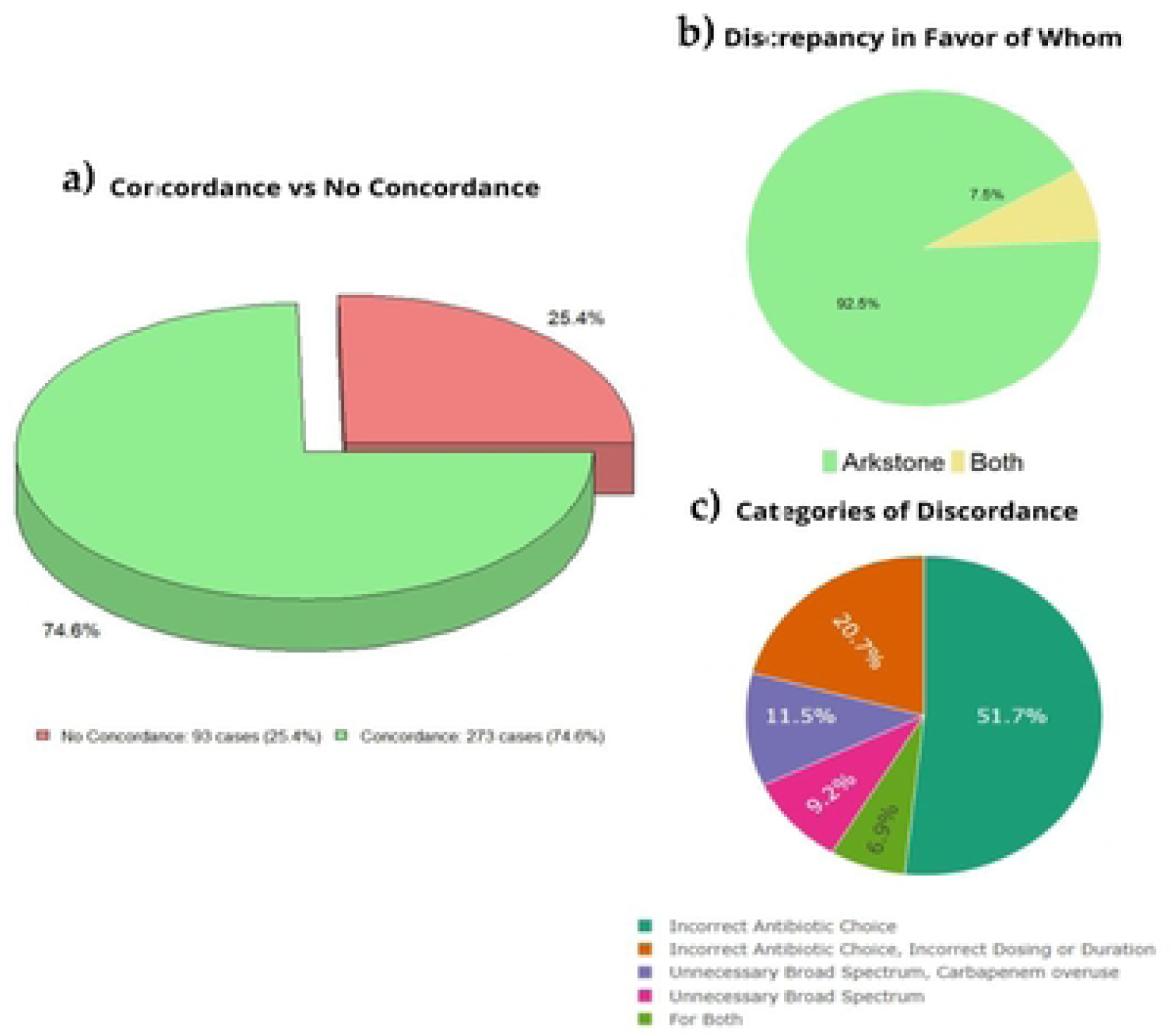
Discordance between CDSS and participants

## 4. Discussion

This study evaluated the concordance between the therapeutic recommendations provided by an artificial intelligence–based clinical decision support system (CDSS), OneChoice®, and expert physician judgment in real-world bacteremia cases. Our findings demonstrate a substantial level of agreement (κ = 0.70), particularly among infectious disease specialists (κ = 0.78), suggesting that the system can offer clinically relevant support when interpreted by trained professionals. These results contribute to the growing body of evidence on the role of CDSS in the management of bloodstream infections (BSIs) and antimicrobial stewardship, particularly in high-resistance settings like Peru [20]. The consistently high antimicrobial resistance rates reported in national surveillance data underscore the urgent need for innovative, effective interventions [20]. The observed lower concordance among non-infectious disease specialists (κ = 0.61) further emphasizes the importance of clinical expertise and reinforces that such tools should serve as supervised decision aids rather than autonomous systems, especially in complex infectious disease contexts.

Our results demonstrate a high concordance (96.14%) between the CDSS used and physician-prescribed treatments, reflecting the potential for AI-driven tools to support antimicrobial decision-making in real-world settings [14]. Substantial agreement (Kappa Index = 0.70) further supports its reliability. These findings align with existing literature highlighting the utility of machine learning-based systems in infectious disease management [12,13].

This study is among the first to evaluate the accuracy of the OneChoice® CDSS in a Peruvian clinical setting and across physician specializations. Higher concordance rates among ID specialists (Kappa = 0.78) compared to non-ID physicians (Kappa = 0.61) highlight the benefit of domain-specific expertise in interpreting CDSS outputs [14]. AI tools such as OneChoice® may be instrumental in bridging the knowledge gap among non-specialist practitioners [12].

Logistic regression identified ID specialization as the primary predictor of concordance (OR = 2.26, p = 0.001), reinforcing the role of targeted training. Interestingly, years of clinical experience showed no significant effect on concordance, suggesting expertise specificity is more relevant than time in practice.

Moreover, the system’s ability to prevent unnecessary carbapenem use in 10.75% of discordant cases aligns with stewardship goals and can reduce selective pressures leading to resistance [11]. Nathan D. Nielsen et al. have noted that CDSS can improve antibiotic appropriateness in critical care settings [22].

This CDSS demonstrated particular strength in cases involving multidrug-resistant (MDR) organisms, such as Pseudomonas aeruginosa, where therapeutic decisions are inherently complex. Notably, P. aeruginosa had the lowest concordance rate (53.84%, p = 0.012), highlighting the critical need for structured tools in guiding antibiotic selection against resistant pathogens [23]. This aligns with prior studies emphasizing that AI-based diagnostics can enhance early pathogen recognition and streamline antimicrobial therapy [24]. While logistic regression analyses identified ID specialization as the strongest predictor of concordance (OR = 2.26, p = 0.001), years of clinical experience did not significantly influence agreement rates. This suggests that expertise in infectious diseases may play a more critical role in aligning with AI-generated recommendations than overall experience [24].

In contrast to the study by Montiel-Romero et al., which reported moderate concordance (κ = 0.48) in antibiotic selection and low concordance (κ = 0.39) in identifying resistance mechanisms between ChatGPT® and infectious disease (ID) specialists in simulated clinical cases [25], our study evaluated an artificial intelligence–based clinical decision support system (OneChoice®) applied to real-world bacteremia cases. We found a higher level of agreement (κ = 0.70) between the system’s recommendations and physician evaluations. Moreover, concordance was significantly higher among ID specialists (κ = 0.78) compared to non-specialists (κ = 0.61). These findings suggest that, unlike general language models such as ChatGPT®, systems specifically trained on structured clinical data may provide more reliable support in real clinical settings.

Nevertheless, consistent with the conclusions of Montiel-Romero et al., our results underscore the importance of using these tools as supervised support rather than as a replacement for clinical judgment, particularly in scenarios where microbiological and pharmacological interpretation is critical for decision-making.

Despite these promising results, the discussion must acknowledge certain limitations. The study’s sample size (366 physician surveys) limits the generalizability of the findings. Also, the retrospective design introduces inherent biases, such as confounders unaccounted for in regression modeling. Additionally, reliance on survey responses could introduce subjective bias, especially if treatment deviated from guidelines [26].

Future research should prioritize prospective, multicenter studies assessing the clinical impact of CDSS on patient outcomes and antibiotic resistance trends. Integration with rapid molecular diagnostics, such as FilmArray® BCID2, could further enhance CDSS utility [27]. Real-time resistance tracking and genomic insights will be critical for refining AI-CDSS applications.

Our findings demonstrate that the CDSS developed by Arkstone provides reliable therapeutic recommendations that align closely with clinical decision-making, particularly among infectious disease specialists. The substantial overall concordance and Kappa Index values indicate that the CDSS is a valuable tool for guiding antimicrobial therapy in bacteremia cases. The higher concordance rates observed among ID specialists and more experienced physicians suggest that specialized training and clinical expertise enhance alignment with CDSS recommendations. Furthermore, the detailed analysis of discrepancies highlights specific areas where the CDSS can aid in optimizing antibiotic selection and dosing, thereby contributing to improved antimicrobial stewardship.

Overall, this CDSS shows significant promise in aligning therapeutic recommendations with best practices, especially for common pathogens and experienced physicians. By reducing variation and enhancing evidence-based decision-making, such tools can support stewardship goals and improve care quality in resource-constrained settings.

## 5. Conclusions

In conclusion, AI-driven CDSS represent a promising advancement in infectious disease management. By enhancing diagnostic precision and standardizing antimicrobial therapy, these systems have the potential to mitigate the global burden of antimicrobial resistance and improve patient outcomes. Future research should build upon these findings to refine and expand CDSS capabilities, ensuring their seamless integration into routine clinical practice.

## Data Availability

The entire database is available in Supplement 4: https://docs.google.com/spreadsheets/d/1gtyjtH8IQGxKfq-2YKXW5fPLHDhoDBEq/edit?gid=685920410#gid=685920410

## Acknowledgments

We thank all the personnel in the Arkstone Medical Solutions and Roe clinical laboratory who have actively been working.

## Notes

### Competing Interest Statement

Ari Frenkel is Chief Science Officer of Arkstone Medical Solutions, the company that produces the OneChoice report evaluated in this study. JC Gómez de la Torre works as the Director of Molecular Informatics at Arkstone Medical Solutions and as the Medical Director at Roe Lab in Perú. At the same time, Alicia Rendon and Miguel Hueda Zavaleta serve as Quality Assurance Managers at Arkstone Medical Solutions. These affiliations may be perceived as potential conflicts of interest. However, the study's design, data collection, analysis, interpretation, manuscript preparation, and the decision to publish the results were conducted independently, with no undue influence from the authors' affiliations or roles within the company.

### Funding Statement

The author(s) received no specific funding for this work.

### Author Declarations

Institutional Ethics Committee of the Private University of Tacna, Peru (FACSA-CEI/093-06-2025).

